# The Charitable Feeding System helps Food Insecure Participants maintain Fruit and Vegetable intake during COVID-19

**DOI:** 10.1101/2021.02.26.21252552

**Authors:** Farryl Bertmann, Katherine Rogomentich, Emily H. Belarmino, Meredith T. Niles

## Abstract

Charitable food services, including food banks and pantries, support individual and households’ food access, potentially maintaining food security and diet quality during emergencies. During the COVID-19 pandemic, the use of food banks and pantries has increased in the US. Here we examine perceptions of the charitable food system and its relationship to food security and dietary quality, specifically fruit and vegetable (FV) intake during the first six months of the COVID-19 pandemic, using a statewide representative survey (n=600) of residents of Vermont. We find that demand for charitable food services increased by 68%. The utilization of food pantries was more common among food insecure households and households with children. Among food insecure respondents, those who used the charitable food system were less likely to reduce their FV intake during the pandemic than those who did not use the charitable food system. Further, we find significant interactions between food pantry use and food insecure households, suggesting that, for food, insecure households, utilizing a food pantry since the onset of the COVID-19 pandemic was associated with higher fruit consumption. These results indicate that these services may support food access and diet quality for at-risk populations during emergencies.

## 1.0 Introduction

The COVID-19 pandemic, associated shutdowns, and social distancing measures designed to slow its spread have profoundly impacted the US food system and food access. According to the Pew Research Center, job disruptions have been widespread; lower-income adults have been hardest hit, with half of their households reporting a job or wage loss due to the pandemic. (1) These disruptions have been disproportionately acute among women, low-income communities, and people of color,(1) which have created significant disruptions to the food supply chain and food security. Recent research suggests that the food insecurity rates have reached levels unprecedented in recent history. (2–4)

With the shift from worksites, schools, and restaurant dining, to greater at-home preparation and consumption, food procurement shifted and, in many cases, overwhelmed grocery stores. (5) Simultaneously, food insecure populations turned to charitable feeding systems (e.g., food banks, pantries). Demands for charitable food services are reported to have increased from 50-140% in the first months of the COVID-19 pandemic. (6,7) By June 2020, nationwide, more than 82% of food banks reported higher numbers of patrons than they did the year prior. (8) A longitudinal population-level survey conducted in Vermont in March and May 2020 found that demand for charitable food services increased by 68%, from 7.1% to 12.0%. (9) In October 2020, Feeding America reported they were on track to distribute 50% more food when comparing October 2019 and October 2020. (10)

Health inequalities in the US follow a socioeconomic continuum where low-income, low-resource households disproportionally experience higher levels of food-related health risks. (11) Further, inequalities, lack of transportation, and geographic disparities magnify structural and environmental factors contributing to food insecurity and poor dietary health.(12,13) Compared to wealthier households, low-income households cook more meals at home (14) yet consume fewer fruits and vegetables (FV)(15) and are more likely not to meet the servings of FV recommended by the Dietary Guidelines for Americans. (16) Nanney et al.(17) examined 269 food shelves using the HEI-2010 (Healthy Eating Index) and concluded that the majority of available food (89%) “needs improvement” for nutritional adequacy. Further, they found significant seasonal fluctuations with the month and quarter scores in July, August, and September significantly higher than in December.

Charitable food services vary in FV distribution from region to region. Vermont is known for its resilient local food system (18) and has several agencies, organizations, and programs to help address hunger issues in the state. According to the Hunger in America 2014(12) Report for Vermont Foodbank, of the 23 meal-based relief agencies analyzed, 42.1% aided clients in accessing local food resources. Further, many sites have introduced client choice(19),(19) to provide food pantry patrons choice; many additional organizations have been transitioning to a client-choice model. This approach allows clients to take products they want and will use. By incorporating behavioral economic techniques, recent initiatives have shown success in nudging clients to select more fruits, vegetables, and nutrient-dense foods. (20) COVID-19 has presented new challenges for these programs as they work to meet growing food needs while protecting staff, volunteers, and clients’ health.

This study aims to understand charitable food programs’ role during the first six months of the COVID-19 pandemic. Emerging international research suggests that COVID mitigation has negatively impacted diet quality during the pandemic. (21) We explore how FV intake changed among a representative sample of Vermonters and examine the emergency food system’s role in maintaining access to FVs during a humanitarian crisis.

## 2.0 Methods

### 2.1 Survey Development and recruitment

The research team, in collaboration with other researchers in the National Food Access and COVID research Team (NFACT)(22), developed a survey in March 2020 (23) with additional refinements in May 2020(9) and August 2020 (24) to measure food access, food security, food purchasing, food assistance program participation, dietary intake, perceptions of COVID-19, and individual social distancing behaviors, as well as household and individual sociodemographics. (22) We obtained Institutional Review Board approval from the University of Vermont (IRB protocol 00000873). The survey was explicitly designed to measure critical outcomes (e.g., food access, food security, food purchasing, and dietary intake) both before the COVID-19 outbreak (dated as of March 11, 2020, the day the World Health Organization declared a global pandemic)(25) and since the pandemic began. The survey utilizes validated measures when possible (Supplementary Table 1). The survey was piloted in Vermont, with 25 eligible (18 or older) residents in late March, and validation methods (e.g., Cronbach alpha, factor analysis) were used to test the internal validity of questions with key constructs (alpha > 0.70). (2)

**Table 1.**
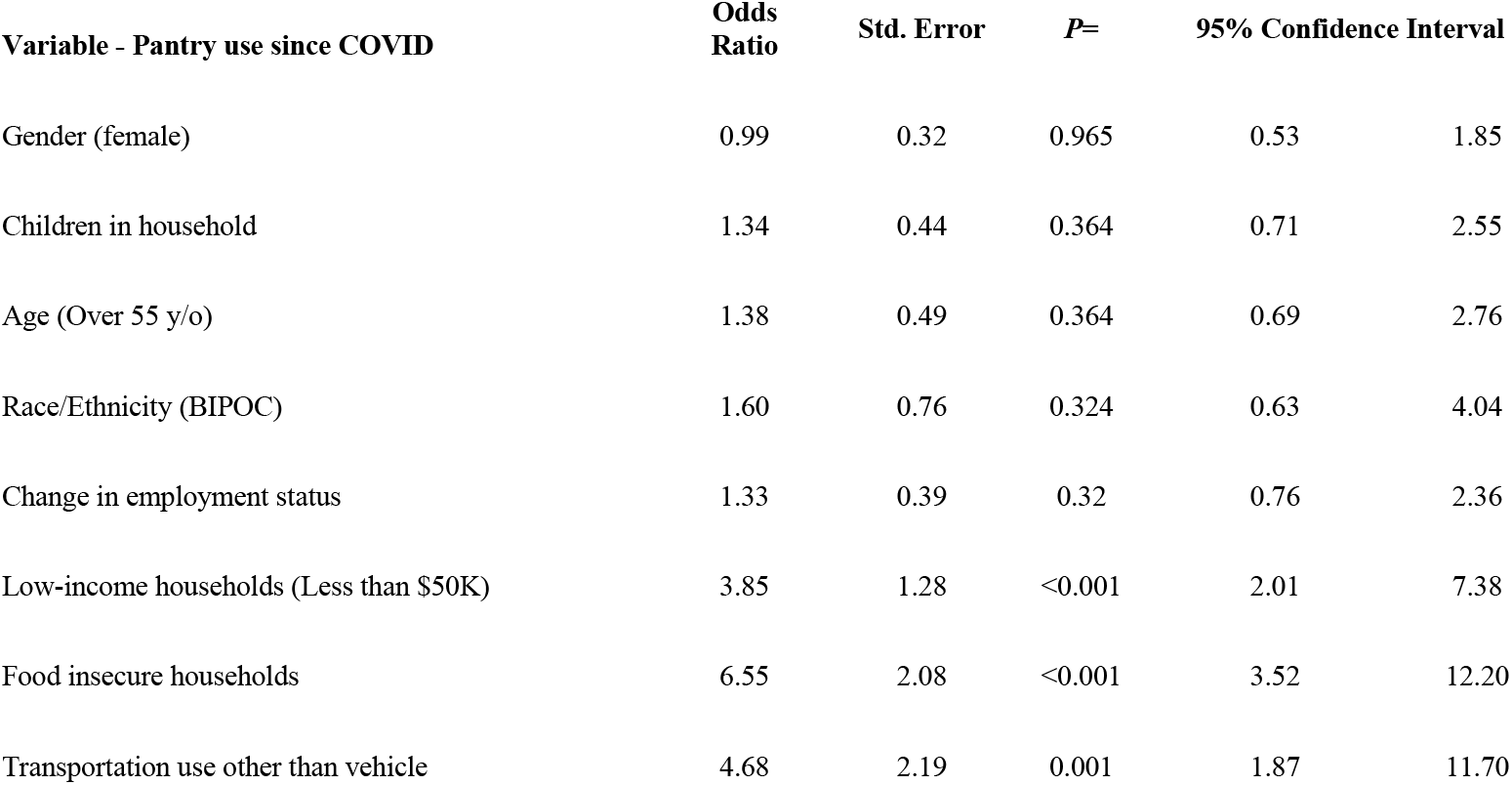
Multivariate analysis predicting odds of food pantry use since the start of the COVID-19 pandemic.

| Variable - Pantry use since COVID | Odds Ratio | Std. Error | P= | 95% Confidence Interval |  |
| --- | --- | --- | --- | --- | --- |
| Gender (female) | 0.99 | 0.32 | 0.965 | 0.53 | 1.85 |
| Children in household | 1.34 | 0.44 | 0.364 | 0.71 | 2.55 |
| Age (Over 55 y/o) | 1.38 | 0.49 | 0.364 | 0.69 | 2.76 |
| Race/Ethnicity (BIPOC) | 1.60 | 0.76 | 0.324 | 0.63 | 4.04 |
| Change in employment status | 1.33 | 0.39 | 0.32 | 0.76 | 2.36 |
| Low-income households (Less than \$50K) | 3.85 | 1.28 | <0.001 | 2.01 | 7.38 |
| Food insecure households | 6.55 | 2.08 | <0.001 | 3.52 | 12.20 |
| Transportation use other than vehicle | 4.68 | 2.19 | 0.001 | 1.87 | 11.70 |

### 2.2 Sampling Approaches

We deployed our online survey to a panel of respondents recruited by Qualtrics (Provo, UT). We developed a sampling strategy for achieving a general population sample representative of the target population for income, race, and ethnicity in Vermont. This sample was achieved by matching sample recruitment quotas to the income, race (specifically White, Black or African American, American Indian and Alaska Native, Asian, Native Hawaiian or Other Pacific Islander, and Two or more races), and ethnicity (Hispanic, non-Hispanic) population profile of Vermont in the American Community Survey (ACS) (Supplementary Table 2). (26) A total of 600 people ages 18 and over responded to the survey, representing a margin of error (95% confidence level) for the adult population of Vermont +/- 4%.(26)

**Table 2.**
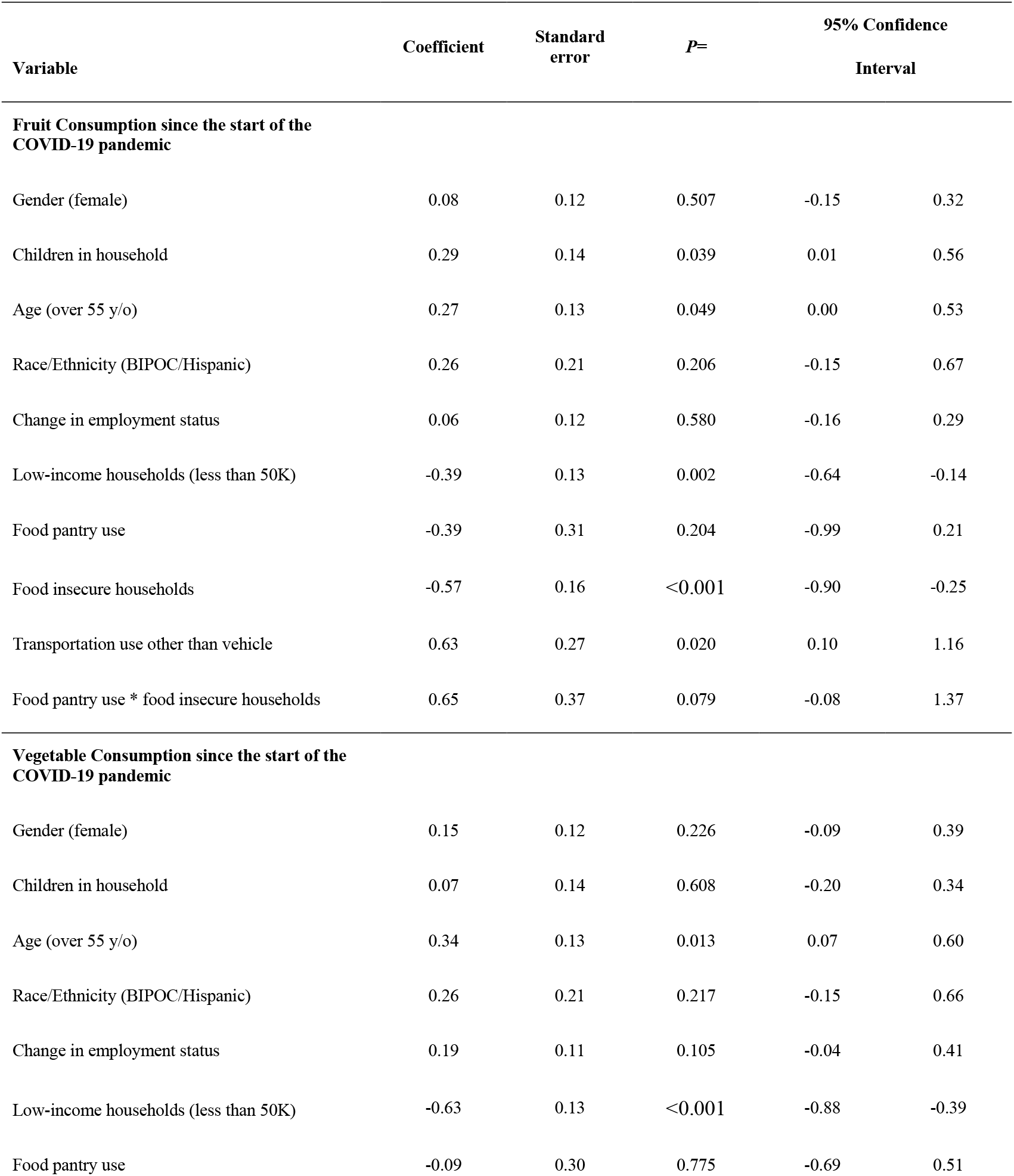
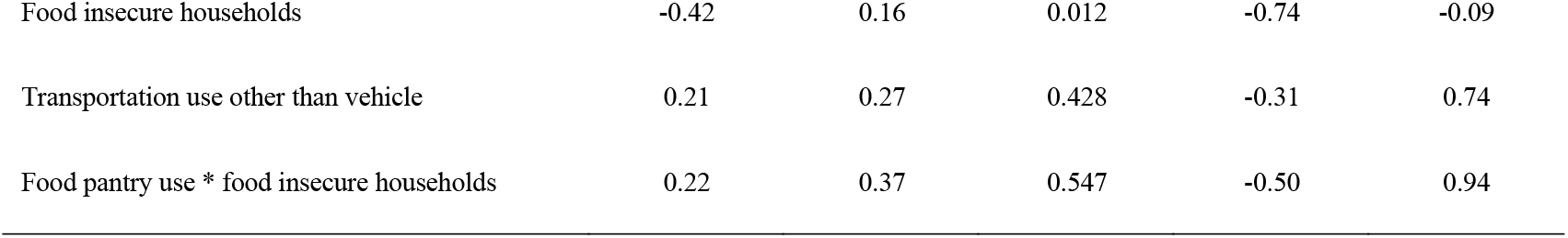
Multivariable regression models predicting fruit and vegetable consumption during the first six months of COVID-19. Models were run independently with separate dependent variables (i.e., fruit consumption and vegetable consumption).

| Variable | Coefficient | Standard error | P= | 95% Confidence Interval |  |
| --- | --- | --- | --- | --- | --- |
| Fruit Consumption since the start of the COVID-19 pandemic |  |  |  |  |  |
| Gender (female) | 0.08 | 0.12 | 0.507 | -0.15 | 0.32 |
| Children in household | 0.29 | 0.14 | 0.039 | 0.01 | 0.56 |
| Age (over 55 y/o) | 0.27 | 0.13 | 0.049 | 0.00 | 0.53 |
| Race/Ethnicity (BIPOC/Hispanic) | 0.26 | 0.21 | 0.206 | -0.15 | 0.67 |
| Change in employment status | 0.06 | 0.12 | 0.580 | -0.16 | 0.29 |
| Low-income households (less than 50K) | -0.39 | 0.13 | 0.002 | -0.64 | -0.14 |
| Food pantry use | -0.39 | 0.31 | 0.204 | -0.99 | 0.21 |
| Food insecure households | -0.57 | 0.16 | <0.001 | -0.90 | -0.25 |
| Transportation use other than vehicle | 0.63 | 0.27 | 0.020 | 0.10 | 1.16 |
| Food pantry use * food insecure households | 0.65 | 0.37 | 0.079 | -0.08 | 1.37 |
| Vegetable Consumption since the start of the COVID-19 pandemic |  |  |  |  |  |
| Gender (female) | 0.15 | 0.12 | 0.226 | -0.09 | 0.39 |
| Children in household | 0.07 | 0.14 | 0.608 | -0.20 | 0.34 |
| Age (over 55 y/o) | 0.34 | 0.13 | 0.013 | 0.07 | 0.60 |
| Race/Ethnicity (BIPOC/Hispanic) | 0.26 | 0.21 | 0.217 | -0.15 | 0.66 |
| Change in employment status | 0.19 | 0.11 | 0.105 | -0.04 | 0.41 |
| Low-income households (less than 50K) | -0.63 | 0.13 | <0.001 | -0.88 | -0.39 |
| Food pantry use | -0.09 | 0.30 | 0.775 | -0.69 | 0.51 |
| Food insecure households | -0.42 | 0.16 | 0.012 | -0.74 | -0.09 |
| Transportation use other than vehicle | 0.21 | 0.27 | 0.428 | -0.31 | 0.74 |
| Food pantry use * food insecure households | 0.22 | 0.37 | 0.547 | -0.50 | 0.94 |

### 2.3 Variables of Interest

We explore three self-reported dependent variables in this analysis (Supplementary Table 1). First, we measured food security status using the US Department of Agriculture’s 6-item short-form food security module. (27) The traditional 12-month period was modified to approximately five months to measure food security status since the start of the COVID-19 pandemic. Following standard scoring protocol, we summarized responses for each item, and classified respondents who answered one or two items affirmatively were categorized as living in food insecure households. Second, we measured current FV intake using the National Cancer Institute’s two-item screener, modified to apply to the last month and with some example foods removed to shorten it. (27) Finally, we examined the perceived change in FV consumption since the onset of the COVID-19 pandemic. Independent variables included multiple questions related to the current food bank and food pantry use, specific charitable food service participant experiences, and transportation other than their own vehicle; we also captured several household and individual-level demographics (Supplementary Table 3).

**Table 3:** Multinomial logit model predicting change in fruit and vegetable consumption during the first six months of the COVID-19 pandemic. The base outcome is no change, so results show the coefficients predicting less or more consumption compared to no change.

| Variable | Coefficient | Standard error | P= | 95% Confidence interval |  |
| --- | --- | --- | --- | --- | --- |
| Less Fruit and Vegetable Consumption since COVID |  |  |  |  |  |
| Gender (female) | 0.26 | 0.27 | 0.329 | -0.27 | 0.80 |
| Children in household | 0.51 | 0.28 | 0.067 | -0.04 | 1.05 |
| Age (over 55 y/o) | -0.13 | 0.29 | 0.664 | -0.70 | 0.44 |
| Race/Ethnicity (BIPOC/Hispanic) | 0.44 | 0.43 | 0.447 | -0.52 | 1.17 |
| Change in employment status | 0.21 | 0.24 | 0.396 | -0.27 | 0.68 |
| Low-income households (less than 50K) | 0.22 | 0.27 | 0.412 | -0.30 | 0.74 |
| Food pantry use | 1.17 | 0.56 | 0.034 | 0.09 | 2.26 |
| Food insecure households | 2.29 | 0.31 | <0.001 | 1.68 | 2.90 |
| Transportation use other than vehicle | -0.23 | 0.49 | 0.637 | -1.20 | 0.73 |
| Food pantry use * food insecure households | -1.74 | 0.66 | 0.008 | -3.03 | -0.45 |
| More Fruit and Vegetable Consumption since COVID |  |  |  |  |  |
| Gender (female) | -0.03 | 0.31 | 0.928 | -0.63 | 0.58 |
| Children in household | 0.44 | 0.34 | 0.201 | -0.23 | 1.11 |
| Age (over 55 y/o) | -0.14 | 0.35 | 0.695 | -0.83 | 0.56 |
| Race/Ethnicity (BIPOC/Hispanic) | 0.96 | 0.43 | 0.026 | 0.12 | 1.79 |
| Change in employment status | 0.55 | 0.30 | 0.063 | -0.03 | 1.13 |
| Low-income households (less than 50K) | -0.46 | 0.33 | 0.172 | -1.11 | 0.20 |
| Food pantry use | 0.68 | 0.71 | 0.337 | -0.71 | 2.08 |
| Food insecure households | 0.70 | 0.45 | 0.122 | -0.19 | 1.58 |
| Transportation use other than vehicle | 0.12 | 0.62 | 0.852 | -1.10 | 1.33 |
| Food pantry use * food insecure households | -0.24 | 0.88 | 0.783 | -1.96 | 1.48 |

### 2.4 Statistical analysis

To examine differences in household food insecurity during the first six months of the COVID-19 pandemic, we created three categories of respondents: 1) *households with food security*, including households that were food secure before and since the onset of the COVID-19 pandemic and households who were food insecure at some point in the year before the COVID-19 pandemic began but were no longer food insecure during the first six months of the pandemic); 2) *households with persistent food insecurity*, food insecure both at some point in the year before the COVID-19 pandemic began and experiencing food insecurity at some point during the first six months of the pandemic during the first six months COVID-19 pandemic); 3) *households with new food insecurity*, categorized as food secure at all times in the year before the COVID-19 pandemic began, but food insecure at some point since the start of the pandemic.

To determine statistically significant differences between groups, we utilized SPSS Version 27(28) and Stata Version 16 (29) to run descriptive statistics, chi-square tests, and multivariable logit models. Specifically, we used chi-square tests to analyze food pantry use related to each item of the food security module. In our multivariable regression models, we use a set of demographic controls including gender, children in the household, respondents over 55, respondents identifying as Black, Indigenous, or People of Color (BIPOC) and/or Hispanic, food security status (27) households with any job loss or negative change since the start of the pandemic, households making less than $50,000 in 2019, and households using transportation for food access other than their own vehicle (e.g. public transportation, ride from a friend) since March 2020. It is important to note that although this survey is representative of Vermont state characteristics on race and ethnicity, the sample size is not sufficient to analyze racial and ethnic groups in a disaggregated format in models. Therefore, we have disaggregated race and ethnicity in all food security statistics in the results but use aggregated race and ethnicity for modeling and matching. We used a multivariable logit model with these demographic controls to predict food pantry use (yes/no) since the start of the COVID-19 pandemic. Then, we use a multinomial logit model with demographic controls to predict a change in FV consumption since COVID-19 (decreased, stayed the same, or increased). Finally, we use a multivariable regression model to predict the current intake of FV, measured on a continuous scale, with demographic controls. All variables and their descriptions are included in Supplementary Table 1. Coefficients are reported in odds ratios for the logistic regression model only. We used all available data to estimate effect sizes and interactions and assumed any missing data were missing at random.

## 3.0 Results

### 3.1 Demographic characteristics of respondents

Our sample reflected the demographic composition of the Vermont population for the gender, race, and income distribution. The majority of our respondents identified as female, non-Hispanic White, without children in the household, and had a household income below $75,000 (Supplementary Table 3). Almost half of the respondents (46.2%) experienced a change in employment at some point between March and September 2020. Changes included loss of employment (24.8%), reduced hours or income (34.7%), and furlough (20.3%). Only 5.0% of respondents utilized transportation other than a personal vehicle between March and September 2020 (Supplementary Table 3).

### 3.2 Food insecurity prevalence

Nearly one in three (29.0%) respondent households were food insecure at some point between March and September 2020. Among those experiencing food insecurity since the start of the pandemic (n=165), 72.1% also experienced food insecurity at some point in the year before the pandemic; in comparison, 27.9% were newly food insecure (Supplementary Table 3).

### 3.3 Fruit and vegetable consumption

The 2020-2025 Dietary Guidelines for Americans (DGA), released on December 28, 2020, recommend that people needing 2,000 calories per day should include at least 2 cups of fruit and 2.5 cups of vegetables in their daily diets. During the COVID-19 pandemic, 15.5% of respondents met the recommendation for fruit intake, and approximately 27.7% of respondents met the recommendations for vegetables (Supplementary Table 3).

### 3.4 Charitable food service utilization

About one in seven respondents (14.5%) reported that their household utilized a food bank or food pantry between March and September 2020 (Supplementary Table 3). Those with increased odds of utilizing charitable food services were food insecure (OR=6.55, 95% CI=3.52, 12.20) and low-income households (OR=3.85, 95% CI=2.01, 7.38), and respondents using transportation other than their own vehicle (OR=4.68, 95% CI=1.87, 11.70) (Supplementary Table 3).

### 3.5 Chartable food service participant experiences

We found that the vast majority of respondents (85%) who utilized food pantries during the first six months of the pandemic (n=86) agreed orlll strongly agreed that food pantries have been helpful (Supplementary Figure 1). Approximately one-third of pantry users indicated concerns, including that pantries run out of food often (35%), have long lines and wait times (34%), and have inconvenient or irregular hours (30%). Other concerns among food pantry users included pantries not having the food their family likes (22%) or good quality food (22%) and not knowing how to prepare food the pantry provides (12%).

**Figure 1:**
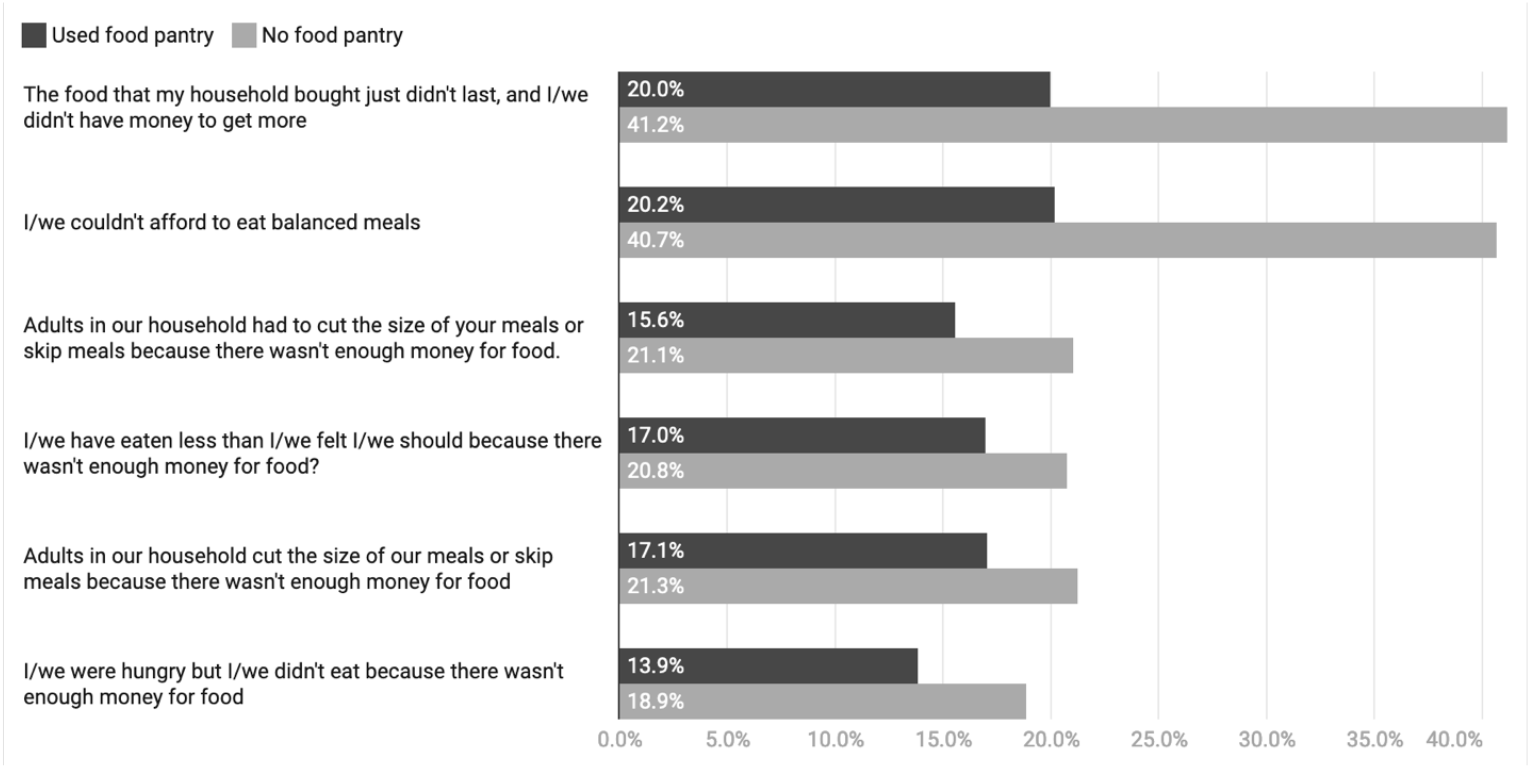
Percent of respondents making less than $50,000 annually that indicated they often or sometimes experienced aspects of food insecurity based on whether or not they used a food pantry since the start of the COVID-19 pandemic (n=259). Chi-squared p-value <0.001 for all differences (Supplementary Table 4).

### 3.6 Pantry utilization buffers aspects of food insecurity among low-income households

Food pantry users were significantly more likely to be food insecure (p<0.001) than non-pantry users. While low-income households (earning less than $50,000 annually) were more likely to use food pantries, we also find that, for low-income households, using food pantries was associated with greater affirmative responses for each food security item (Chi-squared p=<0.001 for all differences (Figure 1)). Expressly, as compared to respondents not using a food pantry, 21% fewer respondents from low-income households who utilized a food pantry since March 2020 agreed that the food they had did not last and they did not have money to get more (20.0%; 41.2%) and that they could afford to eat a balanced meal (20.2%; 40.7%). Among those earning $50,000 annually or less, 60% fewer respondents whose households utilized food pantries agreed that adults in their household had cut the size of their meals or skipped meals because there was not enough money for food as compared to respondents whose households did not utilize food pantries (15.6%; 21.1%). Among the same subset of respondents, four percent fewer respondents whose households utilized the charitable food system reported that they had to eat less (17.0%; 20.8%) or cut the size of their meals or skip meals (17.1%; 21.3%) was not enough money for food.

### 3.7 Fruit and vegetable consumption during the first six months of COVID-19

Using multivariable regression models, we found that respondents in households with children (b=0.29; p = 0.039), those who use a form of transportation other than their own vehicle (b=0.63; p = 0.020), and those over 55 years old (b=0.27; p = 0.049) had higher fruit intake during the first six months of the pandemic than respondents from households without children, those who used their own vehicle, and those aged 18-55 years (Table 2). We found that respondents from low-income households (b=-0.39; p = 0.002) and respondents in food insecure households (b=-0.57; p = 0.001) were more likely to consume less fruit than higher income and food secure households. Furthermore, the interaction of food pantry use and food insecure households was also significant, suggesting that food insecure households utilizing a food pantry since the start of the COVID-19 pandemic was associated with higher fruit consumption (b=0.65; p = 0.079). We found that respondents over 55 years old (b=0.34; p = 0.013) had higher vegetable intake in the first six months of the pandemic compared to younger respondents and those from low-income households (b=-0.63; p = 0.000) and that food insecure households (b=-0.42; p = 0.012) consumed fewer cups of vegetables compared to food secure households.

### 3.8 Changes in Fruit and Vegetable Consumption during the first Six months of COVID-19

Multinomial logit models predicted factors contributing to more, less, or the same FV consumption during the first six months of COVID-19 (p=<0.001, Table 3). Reduced FV consumption was positively associated with having children in the household (b=0.51; p=0.067), food insecure households (b=2.29; p < 0.001), and households utilizing the charitable food system (b=1.17; p = 0.034) since the start of the pandemic. However, respondents who used the charitable food system since the beginning of the COVID-19 pandemic and were food insecure were less likely to reduce their FV intake than respondents who were food insecure and did not use the charitable food system (b=1.74; p = 0.008). Conversely, we found BIPOC/Hispanic respondents were more likely to have increased their FV intake (b=0.96; p = 0.026) during the first six months of the pandemic as compared to non-Hispanic White respondents.

## 4.0 Discussion

To our knowledge, this is the first study to examine the impact of the charitable food system (food banks, pantries/shelves) on FV consumption during the COVID-19 pandemic. Overall, we find that demand for charitable food services increased by 68%, as evidenced by media outlets’ reports. (6) The utilization of food pantries was more common among food insecure households and households with children. Among food insecure respondents, those who used the charitable food system were less likely to reduce their FV intake during the pandemic than those who did not use the charitable food system. Finally, we find an interaction between food pantry use and household food insecurity where food insecure households utilizing a food pantry since the start of the pandemic were significantly associated with higher fruit consumption.

Although low-income households were more likely to prepare home cooked meals before the COVID-19 pandemic,(14) disparities exist in FV intake across socioeconomic status. Home cooked meals are generally associated with higher FV intake. (30) While most households do not eat enough FV – low-income households and those with food insecurity are especially at risk of low FV intake and overall suboptimal diet quality. Higher FV intake is associated with a reduced risk of cardiovascular disease, cancer, co-morbidities, and all-cause mortality. (31) Our results indicate that the charitable food system may play a pivotal role in blunting the adverse effects of a humanitarian crisis like the COVID-19 pandemic on the diet quality of low-income households by providing FV.

Although we found an association between food security status and pantry use, Robaina and Martin (32) demonstrated that our low-income pantry users answered specific statements within the USDA Food Security Module at a significantly lower affirmative rate compared to low-income non-users. We recognize that the USDA defines food security based on Anderson’s 1990 Report(33), where food security is acquired “without resorting to emergency food supplies.” (33) Our findings demonstrate that the charitable food system may have helped maintain several components of food security and FV intake among food insecure users of this system. Our results suggest that although food bank use does not impact the overall food security rate, food security indicators such as “food did not last” and they “could not afford a balanced meal” are associated with positive outcomes among those using food pantries. Further evidence that the charitable food system improves food access includes our findings that 85% of users found food pantries helpful.

As expected, both food insecure and low-income populations are at greater odds of using the charitable food system as compared to food secure and higher income households. We also found the population using any form of transportation other than their own vehicle to be more likely to use the charitable food system, probably due at least in part to the greater reliance on public transportation among low-income persons in the US (34). With state and local social distancing requirements informing distribution, many food pantries have shifted from a super-market-type layout to a drive-up operation where volunteers assembled pre-packaged food boxes and placed them in the patron’s vehicle. (8) Patrons who rely on public transportation may experience barriers to this new food distribution model. Although FV intake did not differ between non-Hispanic White and respondents from racial and ethnic minority populations at the time of our survey, BIPOC/Hispanic respondents were more likely to report a significant increase in FV intake during the first six months of the COVID-19 pandemic. This is notable and important since increasing FV intake is a national public health goal, and FV intake tends to be lower among some racial and ethnic groups. (35) The FV intake among BIPOC respondents mirrors findings in France by Marty et al. (2021), who found an increase in FV consumption during the lockdown. However, their subjects also increased their consumption of sugary foods, sodium, and alcoholic beverage, which our study did not capture. (36)

Prior research by Simmet et al. suggests that the mean number of FV servings provided by food pantries in the US is adequate for the intended number of days. (37) FV distribution varies across different charitable food systems, and often, FVs consisted of tomato sauce, canned vegetables,(37) fruit, and juices. Future inquiry would benefit from mapping regional differences in the form of FV distributed and the origin of these products. We acknowledge that charitable food services are part of a broader system of food access and food security. The charitable food system is designed as an emergency stop-gap and is valuable in crises like the one presented by the COVID-19 pandemic, but does not replace the central role of federal nutrition assistance programs, which are purposely designed to supplement the diverse needs of the most vulnerable Americans. Researchers(38) indicate that the chronic reliance on charitable food services can worsen food security for many households and limit access to culturally and medically appropriate foods. An additional important role of the charitable food system is to help link people to other programs in times of need. It remains to be investigated the extent to which this occurred during the COVID-19 pandemic.

## 4.1 Limitations

We note a few limitations. First, although our approach’s strength was the use of quota sampling to achieve alignment between the sample and the population of Vermont with respect to race, ethnicity, and income, respondents may have differed in other ways. Prior work has demonstrated differences between participants in online survey research and the general population, including greater participation among women, which we saw in our sample.(39,40) Second, self-reported dietary data are subject to recall and response bias. (41) Although the two-item FV intake instrument that we used has adequate reliability, it has low validity for measuring precise intake levels. (42) We used this instrument to compare individuals concerning FV intake rather than estimate actual intake in line with recommendations. (42) Finally, these cross-sectional data do not allow rigorous evaluation of a causal link between food pantry use and food security or dietary quality. Future research should address these limitations and consider the longer-term associations between food pantry use, food security, and diet quality in crisis contexts.

### 4.2 Conclusion

This study documented use and experiences with the charitable food system, including associations with food security and FV intake outcomes, among a statewide sample in Vermont, US, in the first six months of the COVID-19 pandemic. We found that the charitable food system’s use significantly increased in Vermont since the start of the COVID-19 pandemic. The results document improved dietary quality outcomes among low-income households that utilized food pantries as compared with low-income households that did not. Taken together, the results suggest that the charitable food system is an important way in which people can supplement their food budget and maintain food access during a humanitarian crisis. However, it is essential to note that Vermont’s resilient food system and support programs may have impacted these results and the seasonal abundance when this survey was conducted. Additional research should be conducted more fully to understand these relationships over time and in greater depth. Increased analysis of food pantries’ dietary quality in serving diverse populations may be important to assess the overall accessibility of healthy and culturally and medically acceptable foods for at-risk populations. The heightened usage of the charitable food system during the COVID-19 pandemic highlights not only the importance of food pantries but reinforces the need for funding, maintenance, and preparedness of these emergency programs.

## Data Availability

The survey instrument materials used for this current study are available at Harvard Dataverse at: https://dataverse.harvard.edu/dataverse/foodaccessandcoronavirus . The datasets used and/or analyzed during the current study are available from the corresponding author on reasonable request.

## Acknowledgments

Funding for this work was provided by the University of Vermont’s College of Agriculture and Life Sciences and Office of the Vice President of Research, the Gund Institute for Environment at the University of Vermont, and the USDA Agricultural Research Service Center for Food Systems Research.

## Supplementary Material

### 1 Supplementary Figures and Tables

**1.1 Supplementary Table 1.** Use of dependent and independent variables in statistical analysis.

| Variable | Measurement | Scale |
| --- | --- | --- |
| Food (In)Secure | 6 item food security module from USDA. Statements shown in Figure 2. | Binary variable (Affirmative to 2 or more questions= Food Insecure), 0 or 1 affirmatives= Food Secure |
| Fruit Intake | About how many cups of fruit (including 100% pure fruit juice) do you eat or drink each day? Examples of 1 cup for fruit include 1 small apple, 1 large banana, 1 cup (8 oz.) of 100% juice or canned fruit, or ½ cup of dried fruit. (Yaroch et al. 2012) | 0= None, 1= ½ cup or less, 2= ½ to 1 cup, 3=1–2 cups, 4= 2–3 cups, 5= 3–4 cups, 6= 4 cups or more |
| Vegetable Intake | About how many cups of vegetables (including 100% vegetable juice) do you eat or drink each day? Examples of 1 cup of vegetables include 1 cup of cooked leafy greens, 2 cups of lettuce or raw greens, 12 baby carrots, 1 medium potato, or 1 large raw tomato. (Yaroch et al. 2012) |  |
| Fruit/Vegetable Change | I have been eating more, less, or about the same amount of fruits and vegetables per day. | 1= Less, 2= Same, 3= More |
| Food Pantry Use | Which of the following food assistance programs did your household use in the year before the COVID-19 outbreak, if any, and since the COVID-19 outbreak (March 11)? | 1= Use of "food pantry/food bank" since COVID-19 outbreak, 0= No use of "food pantry/food bank" since COVID-19 outbreak. |
| Shared or Public Transportation Use | What were the typical types of transportation you used to get food for your households, in the year before the COVID-19 outbreak and since the COVID-19 outbreak? Check all that apply | 1= use of bus or other public transit, ride from friend/family/neighbor, ride from taxi, someone brings food to me, walk or bike since the COVID-19 outbreak. 0= no use of these modes |
| Female | Which of the following best describes your gender identity? | 1= Female, 0=Male |
| Children in HH | How many people in the following age groups currently live in your household (including you)? Household includes people currently living within your home, including family and non-family members. | 1= Any children in household, 0= No children in household |
| Over 55 | Please select your age group | 1= Respondent 55 or older, 0= Respondent 55 or younger |
| Race/Ethnicity (BIPOC/Hispanic) | What is your race? Are you of Hispanic, Latino, or Spanish origin? | 1= Respondent identify as Asian, Black or African America, Native America, White, Mixed Race, and/or Hispanic, Latino or Spanish origin, 0= Respondent identifies as white and non-Hispanic, Latino or Spanish origin |
| Any Job Change | Have you or anyone in your household experienced a loss of income or job since the COVID-19 outbreak (March 11)? | 1= Any job change (job loss, reduced hours or income at job, furloughed), 0= No job change |
| Less \$50K | Which of the following best describes your household income range in 2019 before taxes? | 1= Household income below \$50,000 a year, 0= Household income above \$50,000 a year |
| HH Size | How many people in the following age groups currently live in your household (including you)? Household includes people currently living within your home, including family and non-family members. | 1= 1 person, 2= 2 people, 3= 3 people, 4= 4 people, 5= 5 people, 6= 6 people, 7=7 people or more |

**1.2 Supplementary Table 2.** Food insecurity by disaggregated race and ethnicity.

|  | Food Insecurity Rate |  |  | p= (chi2 test) |
| --- | --- | --- | --- | --- |
|  | For Demographic Group | For Outside Demographic Group | Total in Demographic Group |  |
| Asian | 25.0% | 29.1% | 4 | 0.858 |
| Black | 50.0% | 28.7% | 8 | 0.188 |
| Native American | 20.0% | 29.1% | 5 | 0.655 |
| Multiple Race | 33.3% | 28.9% | 21 | 0.066 |
| White | 28.7% | 34.2% | 544 | 0.467 |
| BIPOC/Hispanic | 36.2% | 28.4% | 47 | 0.261 |
| Hispanic | 50.0% | 28.4% | 16 | 0.061 |

**1.3 Supplementary Table 3.** Survey respondents’ individual and household demographic characteristics.

| Characteristic | Respondents (N=600) |
| --- | --- |
| Age - no. (%) |  |
| 18-34 | 153 (25.5) |
| 35-54 | 182 (30.3) |
| 55+ | 263 (43.8) |
| Children in household - no. (%) |  |
| Yes | 178 (30.2) |
| No | 415 (70.0) |
| Gender - no. (%) |  |
| Female | 404 (67.3) |
| Male | 190 (31.7) |
| Transgender/Non-binary/Self-Described | 6 (1.0) |
| BIPOC -Race - no. (%) |  |
| White | 559 (93.2) |
| Two or more races | 22 (3.7) |
| American Indian or Alaska Native | 5 (0.8) |
| Asian | 4 (0.7) |
| Black or African American | 9 (1.5) |
| BIPOC - Ethnicity - no. (%) |  |
| Not Hispanic or Latino | 583 (97.2) |
| Hispanic or Latino | 17 (2.8) |
| 2019 Household Income - no. (%) |  |
| Less than \$10,000 per year | 39 (6.5) |
| \$10,000-\$24,999 | 81(13.5) |
| \$25,000-\$49,999 | 141 (23.5) |
| \$50,000-\$74,999 | 110 (18.3) |
| \$75,000 - \$99,999 | 77 (12.8) |
| \$100,000 or more | 145 (24.1) |
| Job change during the COVID-19 pandemic - no. (%) |  |
| Lost job | 149 (24.8) |
| Reduced hours or income | 208 (34.7) |
| Furloughed | 122 (20.3) |
| Any change | 270 (46.23) |
| No changes | 314 (53.8) |
| Food security status during the COVID-19 pandemic - no. (%) |  |
| Food secure | 414 (69.0) |
| Persistently food insecure | 116 (19.3) |
| Newly food insecure | 49 (8.2) |
| Transportation use other than vehicle during the COVID-19 pandemic - no. (%) |  |
| Yes | 30 (5.0) |
| No | 568 (95.0) |
| Daily fruit consumption during the COVID-19 pandemic- no. (%) |  |
| None | 66 (11.0) |
| 1/2 cup or less | 127 (21.2) |
| 1/2 to 1 cup | 158 (26.3) |
| 1-2 cups | 156 (26.0) |
| 2-3 cups | 66 (11.0) |
| 3-4 cups | 15 (2.5) |
| 4 or more cups | 12 (2.0) |
| Daily vegetable consumption during the COVID-19 pandemic- no. (%) |  |
| None | 32 (5.3) |
| 1/2 cup or less | 82 (13.7) |
| 1/2 to 1 cup | 134 (22.3) |
| 1-2 cups | 186 (31.0) |
| 2-3 cups | 106 (17.7) |
| 3-4 cups | 44 (7.3) |
| 4 or more cups | 16 (2.7) |
| Food pantry use during the COVID-19 pandemic - no. (%) |  |
| Yes | 86 (14.5) |
| No | 508 (85.5) |

**1.4 Supplementary Table 4.**
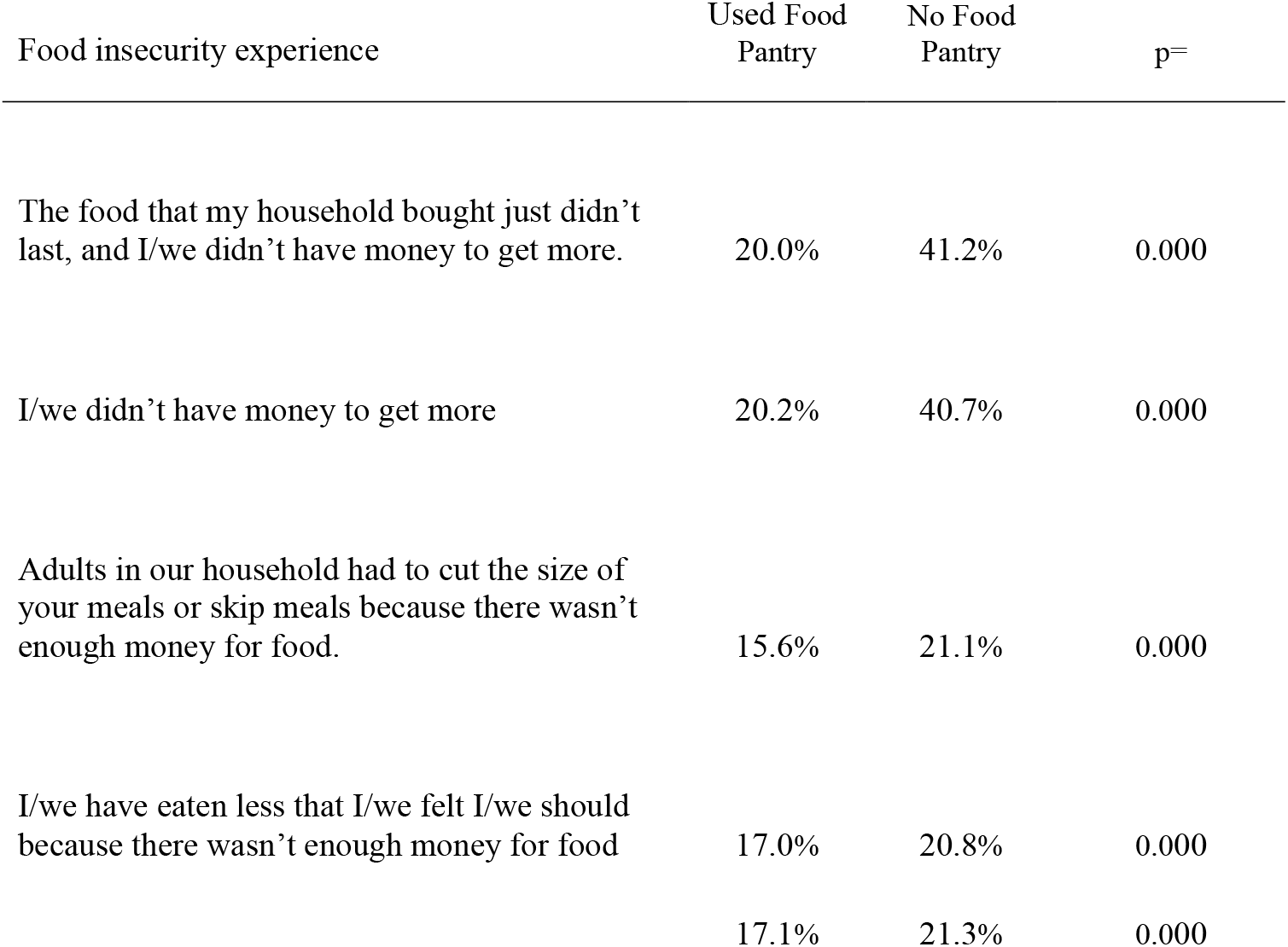
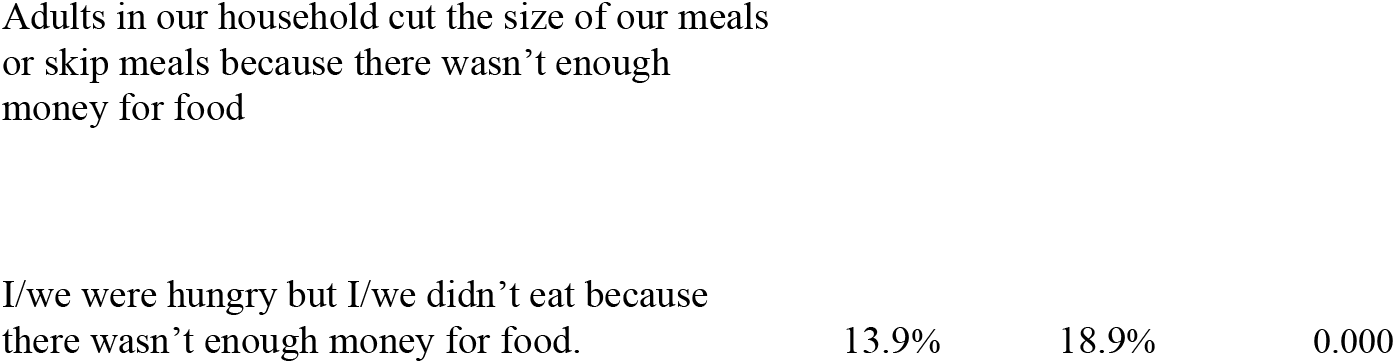
Percent of respondents making less than $50,000 annually that indicated they often or sometimes experienced aspects of food insecurity based on whether or not they used a food pantry since the start of the COVID-19 pandemic (n=259). P values determined through chi-square tests.

| Food insecurity experience | Used Food Pantry | No Food Pantry | p= |
| --- | --- | --- | --- |
| The food that my household bought just didn't last, and I/we didn't have money to get more. | 20.0% | 41.2% | 0.000 |
| I/we didn't have money to get more | 20.2% | 40.7% | 0.000 |
| Adults in our household had to cut the size of your meals or skip meals because there wasn't enough money for food. | 15.6% | 21.1% | 0.000 |
| I/we have eaten less than I/we felt I/we should because there wasn't enough money for food | 17.0% | 20.8% | 0.000 |
|  | 17.1% | 21.3% | 0.000 |
| Adults in our household cut the size of our meals or skip meals because there wasn't enough money for food |  |  |  |
| I/we were hungry but I/we didn't eat because there wasn't enough money for food. | 13.9% | 18.9% | 0.000 |

### 2.0 Supplementary Figures

**Supplementary Figure 1.**
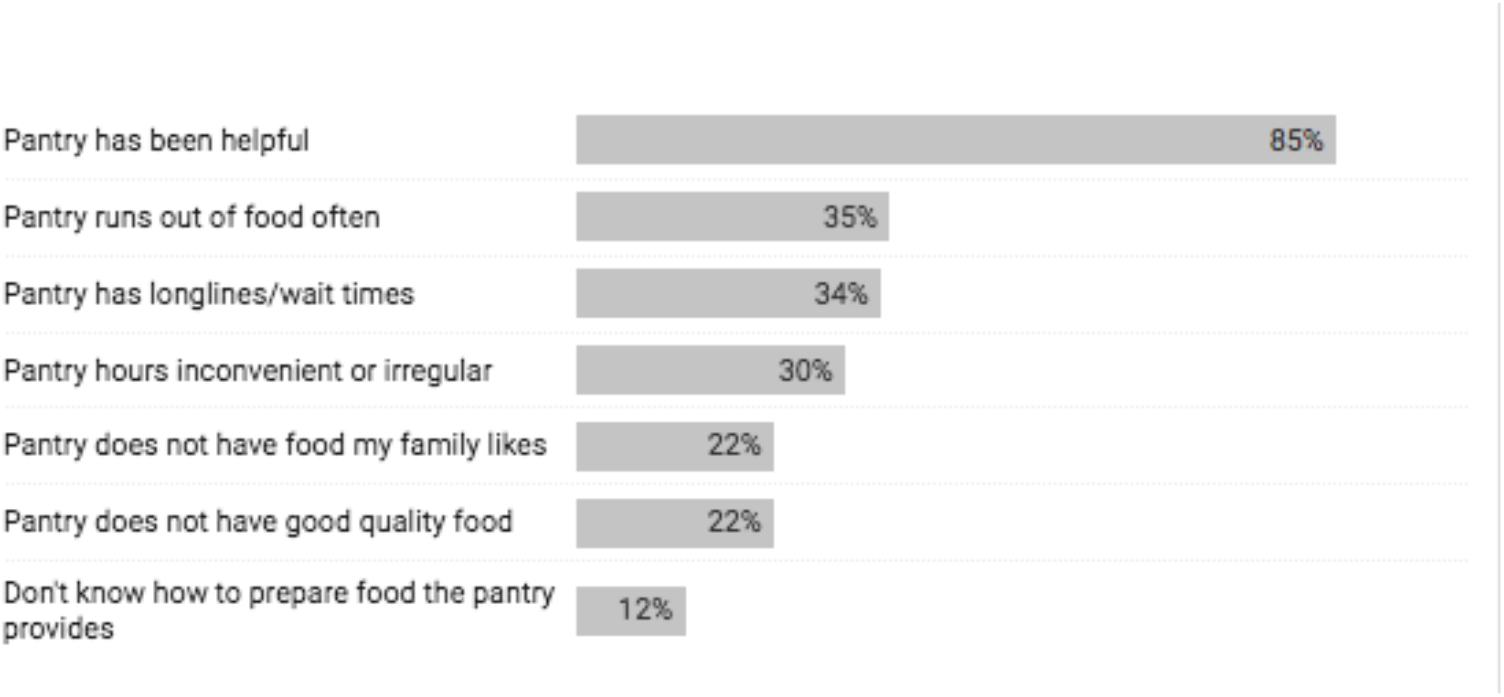
Participant experiences related to using food pantries during the COVID-19 pandemic (N=86). Footnote: Includes respondents who strongly agreed or agreed with the statement

## 11 Data Availability Statement

Availability of data and materials: The survey instrument materials used for this current study are available at Harvard Dataverse at: https://dataverse.harvard.edu/dataverse/foodaccessandcoronavirus. The datasets used and/or analyzed during the current study are available from the corresponding author on reasonable request.

## 12 Declarations

Ethics approval and consent to participate: Institutional Review Board approval was obtained from the University of Vermont under protocol 00000873. Consent was obtained from all participants prior to data collection.

